# Can We Compare Attributable Risk of Adverse Events with the Self-Controlled Case Series Design in Vaccine Safety Studies? A Use Case of Guillain-Barré Syndrome

**DOI:** 10.1101/2025.05.07.25327155

**Authors:** Caihua Liang, Erica L. Chilson, Joanne Wu, Scott P. Kelly, Qing Liu, Kathleen L. Dooling, Maria Maddalena Lino, Joseph A. Lewnard, Bradford D. Gessner, Elizabeth Begier

## Abstract

**Background:** Estimating attributable risk (AR) through self-controlled case series (SCCS) analyses alone may limit generalizability because SCCS only incorporate vaccinated patients with the outcome (e.g., Guillain-Barré syndrome [GBS]) who may differ from the overall population recommended for vaccination.

**Objective:** We aimed to demonstrate background event incidence rates’ impact on vaccine-specific GBS ARs and to standardize ARs across different vaccine studies by applying a generalizable, population-based GBS background rate for improved comparability and to better estimate the expected population-level ARs.

**Methods:** We identified post-licensure SCCS vaccine studies and GBS background rates using US Medicare data via targeted literature review. GBS control period rates from SCCS vaccine studies were extracted or calculated. Population-level ARs were calculated for each vaccine using published background GBS rates and the original SCCS-generated incidence rate ratios (IRRs).

**Results:** Published vaccine-specific GBS IRRs ranged from 2.02 (95%CI: 0.93–4.40) for RSVPreF to 4.96 (95%CI: 1.43–17.27) for recombinant zoster vaccine (RZV). Study-specific ARs per 100,000 doses ranged from 0.28 (H1N1) to 0.90 (RSVPreF). Vaccines with lower control period GBS rates had a lower attributable risk for a given IRR. After standardization using published background GBS rates, population-level ARs were higher for vaccines with higher IRRs. For example, when using the H1N1 vaccine control period rate as the GBS background rate for AR calculation, RSVPreF had the lowest AR per 100,000 doses (0.21) and RZV the highest (2.37).

**Conclusion:** AR calculation is dependent on the control period rate within a given SCCS analysis. ARs derived exclusively from SCCS analyses may lead to incorrect conclusions regarding an adverse event’s absolute risk if included in product labels due to a lack of external validity. Using a representative background rate from the recommended vaccinee population, along with SCCS-derived IRR, to calculate the expected population-level AR may offer more accurate vaccine risk-benefit assessments.

## 1. Introduction

Identification of adverse events that are associated with vaccination versus those which are coincidental and unrelated is important to accurately characterize any risks of vaccination. Rare adverse events may only be detected post-licensure when the vaccine is administered in larger populations [1, 2], highlighting the importance of post-marketing safety surveillance of vaccines. In the US, post-marketing vaccine safety studies often leverage large-scale, real-world data sources, such as Medicare and commercial insurance databases, to evaluate adverse events of special interest, such as Guillain-Barré syndrome (GBS), a rare and severe immune system disorder affecting the peripheral nervous system [3].

The self-controlled case series (SCCS) study design, which is widely used for vaccine safety assessments, compares the incidence of an adverse event (e.g. GBS) during a predefined risk period following exposure (e.g. vaccination) to the control period within the same group of individuals (**Figure 1**). Since the same individuals contribute to both risk and control follow-up time, the primary advantage of SCCS analysis is that it automatically adjusts for time-invariant confounders (e.g., demographics, chronic conditions, health-seeking behaviors) [1, 2, 4]. However, the SCCS methodology has the limitation that it only provides a valid estimate of the relative incidence (e.g., incidence rate ratio [IRR]) but may not provide a representative absolute risk (e.g., attributable risk) of an adverse event [1, 5], i.e., the study-specific attributable risk may lack external validity.

**Fig. 1.**
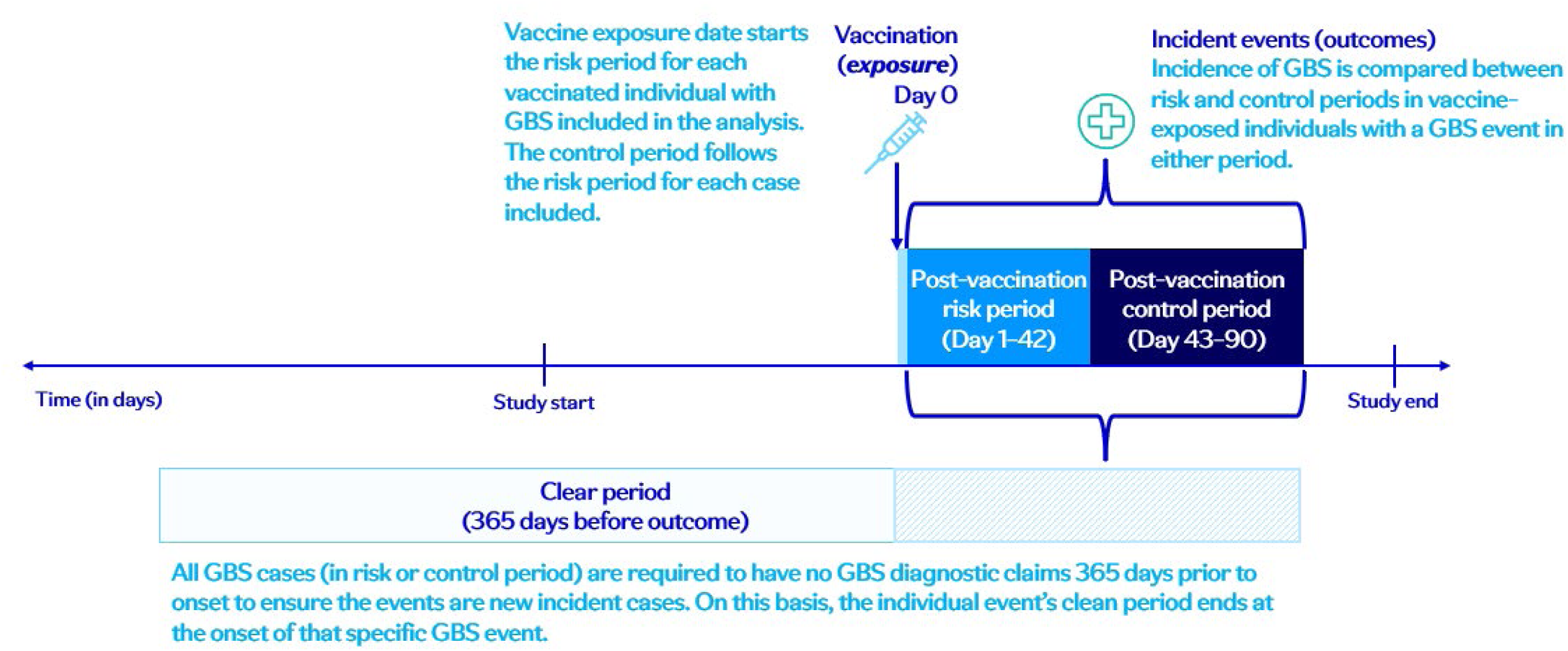
Schematic representation of an example of the self-controlled case series study design for a single dose vaccine*. *This schematic, adapted from the Lloyd et al SCCS analysis [7], may be applied to the safety assessment of any single dose vaccine. The duration of the clear period may vary depending on the occurrence of the incident event (outcome), and the duration of the control period may vary based on the vaccine.

Attributable risk is an estimate of the absolute increase in an event associated with an intervention (e.g., four adverse events per one million vaccine doses administered are due to a specific vaccine) [6]. Incidence event rates from a background reference population are a key mathematical input for calculating attributable risk [5] and directly impact the calculated attributable risk such that a higher background rate for the same IRR/relative risk results in a higher attributable risk, as outlined in the hypothetical example in **Table 1**.

**Table 1.** A hypothetical example to show the impact of background event incidence rates on the calculation of attributable risk.

| **Vaccine** | **Background GBS**<br>**incidence rate**<br>**per 100,000 PY** | **Incidence Rate**<br>**Ratio** | **Control period**<br>**rate** | **Risk period rate** | **Attributable risk**<br>**(Risk-Control)** | **Attributable risk**<br>**per 100,000 PY** |
| --- | --- | --- | --- | --- | --- | --- |
| A | 1 | 2 | 0.00001 | 0.00002 | 0.00001 | 1 |
| B | 10 | 2 | 0.0001 | 0.0002 | 0.0001 | 10 |

Abbreviations: GBS, Guillain-Barré syndrome; PY, person-years.

The SCCS design uses the control period GBS rate as a proxy for the background GBS incidence rate but the incidence of GBS in this control period may not be representative of a vaccine’s recommended population because: 1) only persons with the exposure (e.g., vaccination) and the outcome (e.g., GBS) in the control or risk period are included [4], 2) the control period is often relatively short (e.g., 48 days [7]) limiting total follow-up time for rare events, and 3) initial vaccinee populations after a vaccine launch may not be representative of the entire vaccine indication. However, attributable risk has often been calculated exclusively based on SCCS analyses [5, 7–9] and these attributable risks have been used to assess the risk-benefit balance of the intervention, make comparisons with other vaccinations, and guide vaccine recommendations [10, 11]. To generate a representative attributable risk, the denominator of the control period rate needs to represent the source population from which the exposed cases arose, along with information for the exposed non-cases, unexposed cases, and unexposed non-cases [4]. As suggested by some experts [1, 2], an external background event incidence rate from the population recommended for vaccination can be used to translate the IRR from a SCCS analysis to an attributable risk in a given SCCS vaccine analysis.

Recently, a SCCS study on GBS risk following respiratory syncytial virus (RSV) vaccination in older adults was conducted using Medicare data [7], in which a contradictory relationship between IRR and attributable risk was observed, i.e., the vaccine with the higher IRR had a lower attributable risk because of the different GBS control period rates for the two RSV vaccines, derived from two separate SCCS populations. In our analysis, we demonstrated the impact of background event incidence rates on vaccine-specific attributable risk estimates in several SCCS vaccine studies using GBS as a use case and illustrate a simple approach to standardize the attributable risks across different SCCS vaccine studies for improved comparability by applying a more generalizable, population-based GBS background incidence rate.

## 2. Methods

### 2.1. Review of the literature on vaccine-specific SCCS analyses and data extraction

A targeted literature review was conducted to identify studies evaluating post-licensure vaccine safety in the US Medicare beneficiary population using the SCCS study design. A systematic search was undertaken in MEDLINE (PubMed) and medRxiv preprint server for articles from 2010 onwards in the English language using the following search string: (“Guillain-Barré Syndrome”[MeSH] OR “GBS”) AND (“Self-Controlled” OR “case-only”) AND (“vaccine”[MeSH] OR “vaccination”) AND (“Medicare” OR “CMS”) and (“attributable risk”). For each eligible post-licensure study identified in the literature search, the number of GBS cases in the risk period, person-time in the risk period, GBS control period rate, and IRR were either extracted or calculated from data in the published articles.

### 2.2. Identification of published GBS background rates

To standardize the attributable risk for each vaccine study selected, published GBS background incidence rates were utilized to contextualize the expected attributable risk of potential vaccine provoked GBS incidence in the population recommended for vaccination, i.e., “population-level attributable risk”. The first published GBS background rate was obtained from a large population-based study funded by the FDA Biologics Effectiveness and Safety (BEST) initiative on adverse events of special interest following COVID-19 vaccination, in which the GBS incidence rate estimated from over 26 million adults aged ≥65 years using Medicare fee-for-service data from 2019 [12], which would be representative of the indicated population for vaccines approved for use in older adults, if a concurrent comparison group is not feasible in an observational study. The second published GBS background rate was obtained from the GBS control period rate in a study evaluating chart-confirmed GBS incidence following H1N1 influenza vaccination using SCCS study design [9]. This study involved over 3 million doses of monovalent H1N1 influenza vaccine administered during the 2009–2010 influenza season to hospitalized Medicare beneficiaries aged ≥65 years enrolled in Parts A and B fee-for-service. Both these published GBS background rates were used in the calculation of population-level attributable risk across different SCCS vaccine studies via a standardization process. The purpose of this standardization (i.e., use of a common background rate across vaccines) is to allow valid comparisons between vaccine attributable risks and to provide a population-level attributable risk that is more representative of the recommended vaccinee population.

### 2.3. Calculation of standardized attributable risk across different SCCS vaccine studies

The attributable risk, defined as the estimated excess GBS cases per 100,000 vaccine doses and per 100,000 person-years, and corresponding 95% confidence intervals (CIs) were recalculated for each vaccine using the two Medicare-derived published GBS background rates mentioned above and the original study-specific SCCS-generated IRRs to generate a population-level attributable risk. The calculations were based on the formula below [5]:

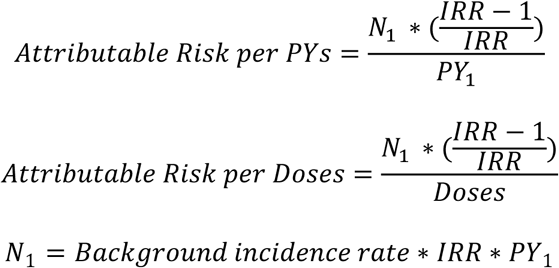

where PY_1_ represents the person-years in the risk period from the SCCS analysis (assuming the same number of individuals who received a vaccine with the same doses and were followed for the same person-time as in the original SCCS studies); and N_1_ represents the updated number of GBS cases in the risk period, given the same IRR and PY_1_ from the original SCCS studies as well as the Medicare-derived published GBS background incidence rate. The 95% confidence intervals for the attributable risks were calculated based on the lower and upper bounds of the IRR 95% CI from the original SCCS studies. All calculations and statistical analyses were performed using R version 4.4.0.

## 3. Results

### 3.1. Vaccine-specific SCCS analyses identified in the literature review

Three vaccine-specific SCCS studies were identified via the targeted literature review as described in section 2.1. Polakowski et al evaluated GBS risk using the self-controlled risk interval study design (i.e., a simplified version of SCCS analysis) in hospitalized Medicare Parts A and B fee-for-service beneficiaries aged ≥65 years who received the 2009 monovalent H1N1 influenza vaccine during the 2009–2010 influenza season [9]. Goud et al estimated the risk of GBS in a SCCS analysis for Medicare Parts A and B fee-for-service and Part D beneficiaries (including inpatient, outpatient and office visits) aged ≥65 years who were vaccinated with recombinant zoster vaccine (RZV) (October 2017 –February 2020) [8]. Lloyd et al compared GBS incidence during a risk period 1–42 days following RSV vaccination (RSVpreF3 or RSVpreF) to the incidence in the control period (43–90 days) in a SCCS design involving Medicare fee-for-service and Part D beneficiaries aged ≥65 years between May 3, 2023–January 28, 2024 [7].

For H1N1 influenza, a GBS control period rate of 1.83 per 100,000 person-years was reported [9]. In case of RZV [8], a GBS control period rate of 0.40 per 100,000 person-years was derived from the reported attributable risk, IRR, and the number of GBS cases using the formula above [5]. Similarly, for the two RSV vaccines, the GBS control period rates of 3.91 (RSVpreF3) and 7.67 (RSVpreF) per 100,000 person-years were calculated using the same method [7].

**Table 2** presents a summary of the three vaccine-specific SCCS analyses based on Medicare beneficiary data. Vaccine-specific GBS IRRs ranged from 2.02 (95% CI: 0.93, 4.40) for RSVpreF [7] to 4.96 (95% CI:1.43, 17.27) for RZV [8]. Published or calculated GBS control period rates per 100,000 person-years ranged from 0.40 (95% CI: 0.11, 1.03) for RZV [8] to 7.67 (95% CI: 3.73, 13.97) for RSVpreF [7]. The corresponding attributable risk of GBS per 100,000 vaccine doses ranged from 0.28 (95% CI: 0.02, 0.55) for H1N1 influenza vaccine [9] to 0.90 (95% CI: −0.02, 1.81) for RSVpreF [7]. Vaccines with lower control period GBS rates were observed to have a lower attributable risk for a given IRR. For example, RZV’s attributable risk of GBS (0.52) was lower than the two RSV vaccines’ attributable risks (0.65 and 0.90) despite a higher IRR of 4.96 because of a lower GBS control period rate of 0.40 per 100,000 person-years.

**Table 2.** Summary of three self-controlled case series vaccine studies using US Medicare data.

| **Vaccine** | **Study period** | **GBS control period rate per 100,000 PY (95% CI)<sup>a</sup>** | **SCCS IRR (95% CI)** | **Attributable risk per 100,000 PY (95% CI)<sup>b</sup>** | **Attributable risk per 100,000 vaccine doses (95% CI)<sup>b</sup>** |
| --- | --- | --- | --- | --- | --- |
| H1N1 influenza vaccine [9] | 2009–2010 | 1.83 (0.95, 3.20) | 2.41 (1.14, 5.11) | 2.47 (0.18, 4.77) | 0.28 (0.02, 0.55) |
| RZV [8] | 2017–2020 | 0.40 (0.11, 1.03) | 4.96 (1.43, 17.27) | 1.59 (0.46, 2.71) | 0.52 (0.15, 0.88) |
| RSVpreF3 [7] | 2023–2024 | 3.91 (1.98, 6.94) | 2.46 (1.19, 5.08) | 5.71 (1.61, 9.80) | 0.65 (0.18, 1.12) |
| RSVpreF [7] | 2023–2024 | 7.67 (3.73, 13.97) | 2.02 (0.93, 4.40) | 7.82 (-0.17, 15.81) | 0.90 (-0.02, 1.81) |

Abbreviations: CI, confidence interval; IRR, incidence rate ratio; PY, person-years; RSV, respiratory syncytial virus; RSVpreF, bivalent (RSV A and B)
stabilized prefusion F protein subunit vaccine; RSVpreF3, the adjuvanted RSV prefusion F protein-based vaccine; RZV, recombinant zoster vaccine; SCCS, self-controlled case series.
<sup>a</sup> GBS control period rate for H1N1 vaccine was obtained from the article; GBS control period rate for RZV was calculated based on the reported attributable risk and IRR using the formula in Section 2.3; GBS control period rates for RSVpreF and RSVpreF3 were calculated based on the reported attributable risk and IRR in the Lloyd et al using the formula in Section 2.3. 95% CIs for control period rates were calculated based on the exact Poisson distribution.
<sup>b</sup> Attributable risk was calculated using the formula in Section 2.3.

### 3.2. Standardizing attributable risk calculation based on published GBS background incidence rates

To calculate the population-level attributable risks across different SCCS vaccine studies for improved comparability, we used the two GBS background incidence rates identified in the published literature. The first published GBS background incidence rate from the FDA BEST initiative study of adverse events of special interest following COVID-19 vaccination was 4.63 cases per 100,000 person-years (95% CI: 4.36, 4.90) [12]. The second published GBS background incidence rate from the H1N1 influenza vaccine study by Polakowski et al was 1.83 cases per 100,000 person-years (95% CI: 0.95, 3.20) [9]. These two published GBS background incidence rates and the SCCS-generated IRRs were used to recalculate the attributable risks (**Table 3**). After standardization, higher attributable risks were observed for those vaccines that had higher IRRs. Standardizing using the published GBS background incidence rate of 4.63 per 100,000 person-years from the Moll et al study [12] decreased the attributable risk per 100,000 vaccine doses from 0.90 (95% CI: −0.02, 1.81) to 0.54 (95% CI: −0.08, 0.83) for RSVpreF and increased the attributable risk from 0.28 (95% CI: 0.02, 0.55) to 0.75 (95% CI: 0.16, 1.03) for H1N1 influenza vaccine, from 0.52 (95% CI: 0.15, 0.88) to 6.00 (95% CI: 2.26, 7.08) for RZV, and from 0.65 (95% CI: 0.18, 1.12) to 0.78 (95% CI: 0.21, 1.05) for RSVpreF3. After standardizing using the published GBS background incidence rate of 1.83 per 100,000 person-years from the H1N1 influenza vaccine SCCS study [9], the attributable risk of GBS per 100,000 vaccine doses decreased from 0.90 (95% CI: −0.02, 1.81) to 0.21 (95% CI: −0.03, 0.33) for RSVpreF and from 0.65 (95% CI: 0.18, 1.12) to 0.31 (95% CI: 0.08, 0.42) for RSVpreF3, while the attributable risk of GBS for RZV increased from 0.52 (95% CI: 0.15, 0.88) to 2.37 (95% CI: 0.89, 2.80). Similar patterns were observed for the attributable risk per 100,000 person-years.

**Table 3.** Standardized population-level attributable risks calculated using published GBS background incidence rates and study-specific IRRs to improve comparability across different SCCS vaccine studies and attributable risk generalizability.

| Vaccine | Study population | Study period | Published GBS background incidence rate per 100,000 PY (95% CI) | Study-specific IRR from SCCS analysis (95% CI) | Population-level attributable risk per 100,000 PY (95% CI) | Population-level attributable risk per 100,000 vaccine doses (95% CI) |
| --- | --- | --- | --- | --- | --- | --- |
| *Utilize GBS background rate from Moll et al., 2023 [12] [Data Years: 2019 to 2020]* |  |  |  |  |  |  |
| H1N1 influenza vaccine [9] | Medicare FFS beneficiaries aged ≥65 years | 2009–2010 | 4.63 (4.36, 4.90) | 2.41 (1.14, 5.11) | 6.53 (1.37, 8.97) | 0.75 (0.16, 1.03) |
| RZV [8] | Medicare FFS and Part D beneficiaries aged ≥65 years | 2017–2020 | 4.63 (4.36, 4.90) | 4.96 (1.43, 17.27) | 18.33 (6.91, 21.64) | 6.00 (2.26, 7.08) |
| RSVpreF3 [7] | Medicare FFS and Part D beneficiaries aged ≥65 years | 2023–2024 | 4.63 (4.36, 4.90) | 2.46 (1.19, 5.08) | 6.76 (1.82, 9.15) | 0.78 (0.21, 1.05) |
| RSVpreF [7] | Medicare FFS and Part D beneficiaries aged ≥65 years | 2023–2024 | 4.63 (4.36, 4.90) | 2.02 (0.93, 4.40) | 4.72 (-0.70, 7.23) | 0.54 (-0.08, 0.83) |
| *Utilize GBS background rate from Polakowski et al., 2013 [9] [Data Years: 2009 to 2010]* |  |  |  |  |  |  |
| H1N1 influenza vaccine [9] | Medicare FFS beneficiaries aged ≥65 years | 2009–2010 | 1.83 (0.95, 3.20) | 2.41 (1.14, 5.11) | 2.47 (0.18, 4.77) | 0.28 (0.02, 0.55) |
| RZV [8] | Medicare FFS and Part D beneficiaries aged ≥65 years | 2017–2020 | 1.83 (0.95, 3.20) | 4.96 (1.43, 17.27) | 7.25 (2.73, 8.55) | 2.37 (0.89, 2.80) |
| RSVpreF3 [7] | Medicare FFS and Part D beneficiaries aged ≥65 years | 2023–2024 | 1.83 (0.95, 3.20) | 2.46 (1.19, 5.08) | 2.67 (0.72, 3.62) | 0.31 (0.08, 0.42) |
| RSVpreF [7] | Medicare FFS and Part D beneficiaries aged ≥65 years | 2023–2024 | 1.83 (0.95, 3.20) | 2.02 (0.93, 4.40) | 1.87 (-0.28, 2.86) | 0.21 (-0.03, 0.33) |

Abbreviations: CI, confidence interval; FFS, fee-for-service; IRR, incidence rate ratio; PY, person-years; RSV, respiratory syncytial virus; RSVpreF, bivalent
(RSV A and B) stabilized prefusion F protein subunit vaccine; RSVpreF3, the adjuvanted RSV prefusion F protein-based vaccine; RZV, recombinant zoster vaccine; SCCS, self-controlled case series.

## 4. Discussion

Attributable risk is often used to assess the risk-benefit balance of a vaccine, guide vaccine technical committee recommendations, and ultimately help providers communicate the risk to the public [10, 11]. Given these important implications, it is essential to provide an assessment of attributable risk that reflects the recommended vaccinee population. If attributable risk is calculated based only on data from a SCCS analysis, the attributable risk is impacted by the study-specific GBS control period rate, such that vaccines with a lower GBS control period rate have a lower attributable risk for a given IRR. For example, RZV’s attributable risk of 0.52 per 100,000 doses was lower than the two RSV vaccines’ attributable risks of 0.65 (RSVpreF3) and 0.90 (RSVpreF), despite having a higher IRR of 4.96, due to a lower GBS control period rate of 0.40 per 100,000 person-years. Our findings emphasize the non-comparability of attributable risk estimates that are derived from an individual SCCS study where SCCS control period rates of GBS are based on a study population which is both exposed and has the outcome of interest—in other words, a non-generalizable population. Prevalence and incidence rates of GBS in the US have been increasing over time [13]. This is likely contributing in part to the lower attributable risk seen for a much higher IRR for RZV, as the data were collected earlier. Specifically, the United States Prescribing Information (USPI) of RZV utilized an SCCS-based IRR of 4.96 (95% CI: 1.43, 17.27) associated with an “estimated 6 excess cases of GBS per million doses administered to adults aged 65 years or older” (i.e., attributable risk=0.52 per 100,000 doses) [8]. Similarly, the monovalent H1N1 influenza vaccine was reported to have an IRR of 2.41 (95% CI: 1.14, 5.11), with a corresponding attributable risk of 2.84 (0.21, 5.48) per million [9]. These relatively lower attributable risks compared to that of the RSV vaccines are due to the lower control period GBS rates in the RZV and H1N1 influenza vaccinee populations from their respective SCCS vaccine studies. These findings highlight the importance and impact of background event incidence rates on attributable risk calculation and the benefit of using a background rate from a population representative of those recommended for vaccination to standardize to a population-level attributable risk.

Warnings regarding potential increased risk of GBS are present in the prescribing information of several commonly used vaccines [14–18] based on the results from a SCCS study design. The risk of vaccine associated GBS in these warnings is usually expressed as an excess cases per million doses—for example, 1–2 additional cases per one million doses for influenza vaccine [19], and 3–6 additional cases per one million doses for RZV [20]. Recognizing the impact of deriving attributable risk estimates solely from SCCS data, the FDA has included a disclaimer in the recent US package insert update for subunit RSV vaccines: “the background risk of GBS in a study population influences the excess GBS case estimate and may differ between studies, precluding direct comparison to excess GBS case estimates from other vaccine studies or populations” [21, 22]. Their recent study manuscript also includes this caution [7]. However, such SCCS-derived attributable risks have been applied to hypothetical populations of a million future vaccinees and used to compare the risk-benefit balance between vaccines [10].

While calculation of attributable risk aims to help healthcare providers and the public contextualize and interpret risk, if not methodologically sound, it may also lead to confusion and inappropriate comparisons between different vaccine brands [10]. For example, an Advisory Committee on Immunization Practices (ACIP) slide presentation from October 2024 concluded that the potential risk of GBS after protein subunit RSV vaccination was possibly higher than any marketed adult vaccine [11]. However, the IRR for other marketed vaccines is higher than these vaccines. The attributable risk is higher due to higher background risk of GBS in the SCCS study population.

This population-level attributable risk analysis has limitations. First, geographic, demographic, and methodological variations in background rates are a challenge to accurately determining GBS background incidence rates in the population recommended for vaccination. On this basis, the two GBS background incidence rates derived from the US Medicare population may not represent the contemporary population recommended for a specific vaccine because they are from 5 (RZV: 2017–2020) to 15 years (H1N1: 2009–2010) ago. Second, the updated analysis did not consider the potential effect modification with regards to the original IRR from SCCS studies and did not address any potential unmeasured time-varying confounders. Effect modification, if present, may impact the validity of applying a SCCS IRR to an external background GBS rate if the effect modifier is not present in the same frequency in the population from which the external background rate is derived. However, at this time, effect modifiers of the vaccine GBS relationship have not been identified, and we made efforts to harmonize the population characteristics by deriving the background rates from the same database (i.e., Medicare). Therefore, the SCCS-derived population-level attributable risk using the generalizable published background rate could still be impacted by these limitations.

This study raises questions about the inclusion of attributable risk estimates derived exclusively from an SCCS analysis in product labels and direct comparisons of attributable risk across vaccine studies or populations in vaccine risk-benefit assessments. Inaccurate estimation of attributable risk can affect interpretations, clinical decision-making, and recommendations by public health and regulatory agencies.

## 5. Conclusions

Attributable risk calculation is dependent on the background event rate, and in the case of attributable risks derived exclusively from SCCS analyses, a higher control period event rate results in a higher attributable risk. The control period event rate, including that of GBS, is dependent on the characteristics of the vaccinated GBS cases, which may vary across vaccine studies and even among different vaccines within the same study and may not accurately represent the recommended population for the specific vaccine. This raises questions about the inclusion of attributable risk derived exclusively from an SCCS analysis in product labels and comparisons of attributable risk across vaccines, because such study-specific attributable risks may lack external validity and lead to incorrect conclusions about the expected absolute risk of GBS in the broader vaccinee population. To mitigate these issues, it is advisable to use an external GBS background incidence rate from the population recommended for vaccination, instead of the GBS control period rate from the SCCS population, in combination with the SCCS-derived IRR to calculate a population-level attributable risk to allow for more accurate risk-benefit assessments regarding future vaccination and improved comparability across different SCCS vaccine studies. Future studies specific to the populations recommended for vaccination could help establish contemporary background event rates for use in such standardization calculations to ensure the rates used are representative of the current recommended vaccinee population.

## Data Availability

All data produced in the present work are contained in the manuscript

## Abbreviations

CI: confidence interval;
FFS: fee-for-service;
GBS: Guillain-Barré syndrome;
IRR: incidence rate ratio;
PY: person-years;
RSV: respiratory syncytial virus;
RSVpreF: bivalent (RSV A and B) stabilized prefusion F protein subunit vaccine;
RSVpreF3: the adjuvanted RSV prefusion F protein-based vaccine;
RZV: recombinant zoster vaccine;
SCCS: self-controlled case series.

## CRediT authorship contribution statement

**Caihua Liang:** conceptualization; data curation; formal analysis; methodology; software; writing – original draft; writing – review and editing. **Erica L. Chilson:** writing – review and editing. **Joanne Wu:** writing – review and editing. **Scott P. Kelly:** writing – review and editing. **Qing Liu:** formal analysis; software; validation; visualization; writing – review and editing. **Kathleen L. Dooling:** writing – review and editing. **Maria Maddalena Lino:** writing – review and editing. **Joseph A. Lewnard:** methodology; review of formal analysis. **Bradford D. Gessner:** conceptualization; supervision; writing – review and editing. **Elizabeth Begier:** conceptualization; data curation; funding acquisition; investigation; methodology; resources; supervision; visualization; writing – original draft; writing – review and editing. All authors attest they meet the ICMJE criteria for authorship.

## Data availability statement

All data was derived from published studies cited in this manuscript.

## Acknowledgments

Medical writing and editorial assistance for this manuscript was provided by Sudipta Chatterjee and Melissa Furtado, both employees of Pfizer, and funded by Pfizer.

## References

[1] Grosso A, Douglas I, MacAllister R, Petersen I, Smeeth L, Hingorani AD. Use of the self-controlled case series method in drug safety assessment. Expert Opin Drug Saf. 2011;10:337–40. 10.1517/14740338.2011.562187.

[2] Baker MA, Lieu TA, Li L, Hua W, Qiang Y, Kawai AT, et al. A vaccine study design selection framework for the postlicensure rapid immunization safety monitoring program. Am J Epidemiol. 2015;181:608–18. 10.1093/aje/kwu322.

[3] Centers for Disease Control and Prevention. Guillain-Barré Syndrome (GBS) and Vaccines, https://www.cdc.gov/vaccine-safety/about/guillain-barre.html [accessed 7 February 2025].

[4] Eiffert SR, Raman SR. Re: ‘Self-controlled case series design in vaccine safety: a systematic review’ - absolute and relative measures. Expert Rev Vaccines. 2023;22:419-20. 10.1080/14760584.2023.2211165.

[5] Wilson K, Hawken S. Drug safety studies and measures of effect using the self-controlled case series design. Pharmacoepidemiol Drug Saf. 2013;22:108–10. 10.1002/pds.3337.

[6] Szklo M, Nieto FJ. Chapter 3: Measuring Associations Between Exposures and Outcomes. In: Epidemiology: Beyond the Basics. 2nd edition: Jones & Bartlett Publishers; 2007. p. 83-9.

[7] Lloyd PC, Shah PB, Zhang HT, Shah N, Nair N, Wan Z, et al. Evaluation of Guillain-Barré syndrome following Respiratory Syncytial Virus Vaccination among Medicare Beneficiaries 65 Years and Older. medRxiv. 2025. 10.1101/2024.12.27.24319702.

[8] Goud R, Lufkin B, Duffy J, Whitaker B, Wong HL, Liao J, et al. Risk of Guillain-Barré Syndrome Following Recombinant Zoster Vaccine in Medicare Beneficiaries. JAMA Intern Med. 2021;181:1623–30. 10.1001/jamainternmed.2021.6227.

[9] Polakowski LL, Sandhu SK, Martin DB, Ball R, Macurdy TE, Franks RL, et al. Chart-confirmed guillain-barre syndrome after 2009 H1N1 influenza vaccination among the Medicare population, 2009-2010. Am J Epidemiol. 2013;178:962–73. 10.1093/aje/kwt051.

[10] Britton A, Roper LE, Kotton CN, Hutton DW, Fleming-Dutra KE, Godfrey M, et al. Use of Respiratory Syncytial Virus Vaccines in Adults Aged ≥60 Years: Updated Recommendations of the Advisory Committee on Immunization Practices - United States, 2024. MMWR Morb Mortal Wkly Rep. 2024;73:696–702. 10.15585/mmwr.mm7332e1.

[11] Melgar M, Britton A, CDC Advisory Committee on Immunization Practices. RSV Vaccination in Adults: Work Group Interpretations, https://www.cdc.gov/acip/downloads/slides-2024-10-23-24/06-RSV-Adult-Melgar-508.pdf [accessed 28 February 2025].

[12] Moll K, Lufkin B, Fingar KR, Zhou CK, Tworkoski E, Shi C, et al. Background rates of adverse events of special interest for COVID-19 vaccine safety monitoring in the United States, 2019-2020. Vaccine. 2023;41:333–53. 10.1016/j.vaccine.2022.11.003.

[13] Kelly S, et al. Temporal Trends in the Incidence of Guillain-Barré Syndrome in the United States from 2017 to 2023. Manuscript in development.

[14] Food and Drug Administration. Prescribing Information for SHINGRIX (Zoster Vaccine Recombinant, Adjuvanted), suspension for intramuscular injection, 2017 (revised May 2023), https://www.fda.gov/media/108597/download [accessed 28 February 2025].

[15] Food and Drug Administration. Prescribing Information for BOOSTRIX (Tetanus Toxoid, Reduced Diphtheria Toxoid and Acellular Pertussis Vaccine, Adsorbed) injectable suspension, for intramuscular use, 2005 (revised 2023), https://www.fda.gov/media/124002/download [accessed 28 February 2025].

[16] Food and Drug Administration. Prescribing Information for FLUAD^®^ QUADRIVALENT (Influenza Vaccine, Adjuvanted) Injectable Emulsion for Intramuscular Use 2023-2024 Formula, 2020 (revised March 2023), https://www.fda.gov/media/135432/download [accessed 28 February 2025].

[17] Food and Drug Adminstration. Prescribing Information for Adacel (Tetanus Toxoid, Reduced Diphtheria Toxoid and Acellular Pertussis Vaccine Adsorbed), Suspension for Intramuscular Injection, 2005 (revised 2023), https://www.fda.gov/media/119862/download [accessed 28 February 2025].

[18] Food and Drug Adminstration. Prescribing Information for IMOVAX^®^ Rabies Vaccine, 2019, https://www.fda.gov/media/75709/download [accessed 28 February 2025].

[19] Vellozzi C, Iqbal S, Broder K. Guillain-Barre syndrome, influenza, and influenza vaccination: the epidemiologic evidence. Clin Infect Dis. 2014;58:1149–55. 10.1093/cid/ciu005.

[20] Janusz CB, Anderson TC, Leidner AJ, Lee GM, Dooling K, Prosser LA. Projected risks and health benefits of vaccination against herpes zoster and related complications in US adults. Hum Vaccin Immunother. 2022;18:2060668. 10.1080/21645515.2022.2060668.

[21] Pfizer Inc. Prescribing Information for ABRYSVO® (Respiratory Syncytial Virus Vaccine) for injection, for intramuscular use, 2023 (revised January 2025), https://labeling.pfizer.com/ShowLabeling.aspx?id=19589 [accessed 28 February 2025].

[22] GlaxoSmithKline Biologicals. Prescribing Information for AREXVY (Respiratory Syncytial Virus Vaccine, Adjuvanted) for injectable suspension, for intramuscular use, 2023 (revised January 2025), https://gskpro.com/content/dam/global/hcpportal/en_US/Prescribing_Information/Arexvy/pdf/AREXVY.PDF [accessed 28 February 2025].

